# A Test-Based Strategy for Safely Shortening Quarantine for COVID-19

**DOI:** 10.1101/2020.11.24.20238287

**Authors:** Dana M. Lewis, Scott Leibrand, Howard Leibrand

## Abstract

Quarantine for COVID-19 is hard and could safely be made easier. Lengthy quarantine is difficult for many individuals to rigorously observe. To date, 14 days has been recommended as the quarantine period for people who are exposed to a confirmed COVID-19 case, because most infections appear by then. For household close contacts, there remains a 0.5% chance that a COVID-19 infection may appear after that time, because the COVID-19 incubation period is up to 21 days. However, the risk of transmission after 14 days is so small that public health authorities have decided that 0.5% is an acceptable risk to balance the impact of quarantine on individuals, their communities, and our economy. Therefore, this 0.5% risk of infection is a useful benchmark to consider additional strategies for reducing the burden on individuals while reducing spread of COVID-19. With appropriately timed testing and awareness of possible symptoms, a non-symptomatic individual with a negative COVID-19 test at day 7-8 would be at lower risk of being or becoming infectious than someone completing a 14-day quarantine without testing. If we implemented a test-based strategy for determining the end of quarantine, these individuals could be safely released from quarantine at an earlier date, and would be able to more rigorously quarantine themselves while potentially most infectious.

---

The actions we should take to mitigate the risk of getting and transmitting COVID-19 in our communities should be graduated and risk-dependent based on our personal situation, community transmission rates, and more. Some possible actions are easy enough that we ask everyone to do them, like physical distancing, mask wearing, and hand washing. Others, like isolation of those infected and quarantine of those exposed via close contact with a confirmed case, are highly burdensome but necessary on a temporary basis when someone’s transmission risk is far higher.

One key additional piece of information that allows us to better estimate someone’s risk of still being infectious after a given duration of quarantine is the result of a COVID-19 test administered after 7 or more days of quarantine.

To determine the likelihood of being COVID positive even after a certain number of days of symptom-free quarantine, we multiply the proportion of eventually-symptomatic cases showing symptoms by a particular day from Bi et. al.^1^ with a prior probability of infection (such as 10%), and find close agreement with the Reich et. al.^2^ *Undetected Infections* calculator (using data from Zhao et. al.^3^) estimate of *Number of undetected symptomatic infections per 10,000 monitored* using an initial risk of 10% and starting 0 *Days since infectious exposure*. To determine the average probability of being COVID positive even after a certain number of days of symptom-free quarantine followed by a negative test, we multiply the average of the Reich/Zhao and Bi quarantine-only estimates by the Kucirka et. al.^4^ probability of someone COVID positive testing PCR negative after a certain number of days.

By this method, we can calculate that an individual with a 10% prior probability of infection who has a negative test on day 8 has a posttest probability of being infectious of 0.4%, which is slightly less than the probability of such an individual becoming infectious when exiting quarantine after 14 days without a test.

This relationship holds regardless of someone’s initial exposure risk, and likely regardless of the type of COVID-19 test they receive: they are slightly less likely to remain infectious after 8 days with a negative test than after 14 days without a test.

**Figure 1:**
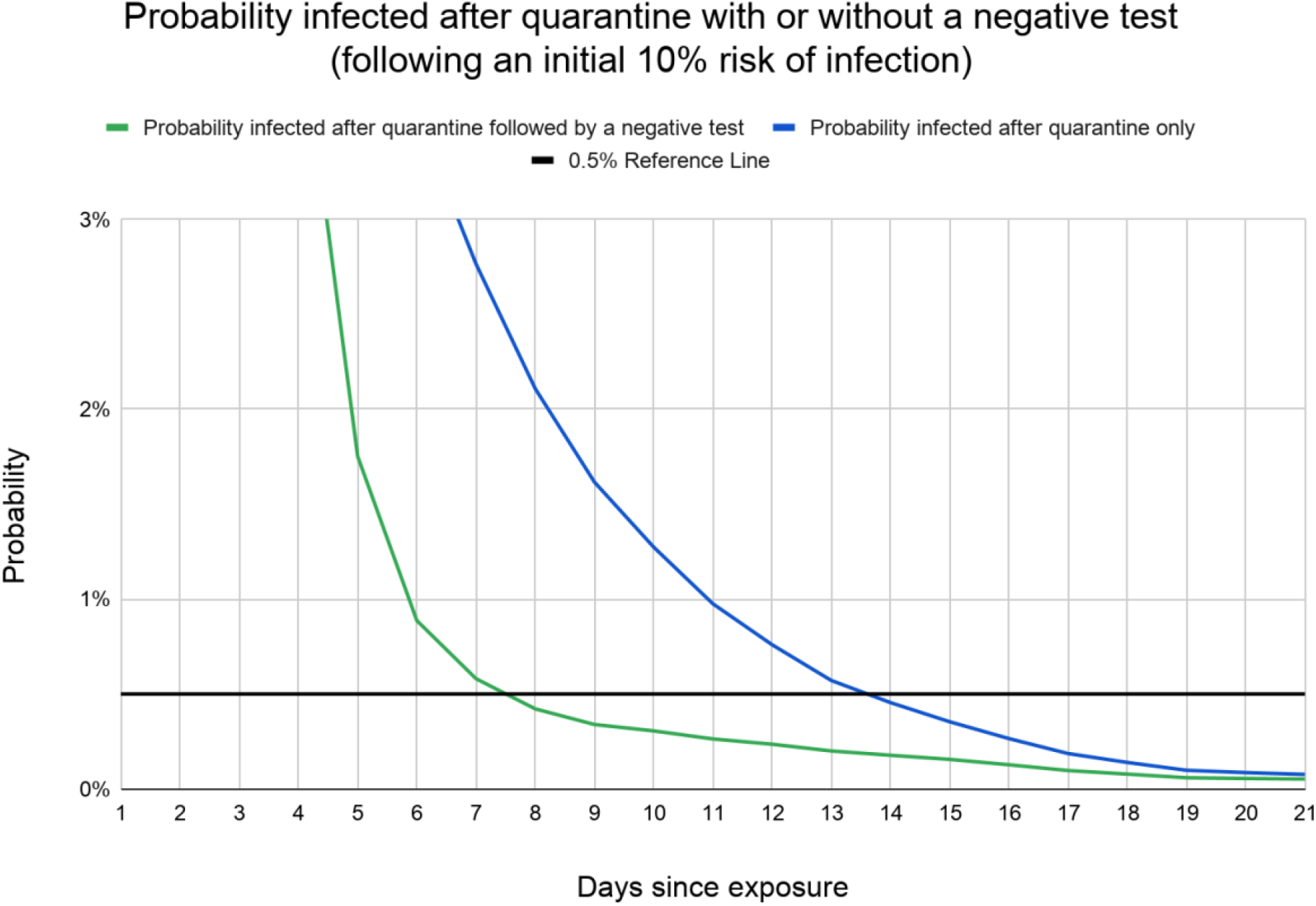
The risk of remaining infectious drops to 1/20th of the initial exposure risk (from 10% to 0.5% in this example) after 14 days of quarantine without development of symptoms. With quarantine followed by a negative test, the same risk reduction is achieved by day 8.

Because even a highly sensitive test like PCR, if given only once, has a significant chance of missing an infection, quarantine guidelines have previously avoided depending on such information. However, some public health guidelines are now beginning to recognize this equivalence of 14-day quarantines and shorter quarantines accompanied by a negative test. For example, Washington State’s guidance issued beginning November 16^5^ recommends that indoor social gatherings be avoided except among individuals who have quarantined for 14 days, or for 7 days accompanied by a negative test. No matter what someone’s baseline level of risk is, that risk can be equivalently reduced by either approach.

Another benefit of mid-quarantine testing is that it allows for earlier (and likely stricter) self-isolation of those who test positive, and earlier quarantine of any contacts they might have exposed. It also simplifies quarantine management, as public health officials needn’t worry about individuals deciding to leave quarantine after receiving a negative test result, provided they’re tested at least 7 days after potential exposure.

Public health officials often make hard choices about what can be done to encourage behaviors that move public health in the intended direction. Some officials may have discouraged testing during quarantine, fearing that individuals with a negative test may stop or loosen quarantine, reducing its efficacy. But with a testing-based strategy, public health officials can encourage testing at the most ideal time. This both encourages the behavior needed by the public and minimizes the burden on individuals, hopefully improving quarantine compliance overall.

In summary, we can make quarantine a more effective and tolerable tool by adding testing after day 7. Negative tests can allow for early exit from quarantine without increased risk. Tests that come back positive can help us identify asymptomatic infected individuals earlier and thus manage them and their contacts more effectively. Educating quarantined individuals about the best timing for testing and the fact that a properly timed negative test would end their quarantine may encourage them to more rigorously quarantine themselves while potentially most infectious.

--

**Dana Lewis** and **Scott Leibrand** are volunteer consultants to public health officials in Washington state and co-invented the first open source Bluetooth exposure notification protocol for public health applications. **Dr. Howard Leibrand** is the Skagit County Health Officer in Washington state and co-author of the MMWR article “*High SARS-CoV-2 Attack Rate Following Exposure at a Choir Practice”*.

## Supporting information

Supplementary data Calculating Probability infected after quarantine with or without a negative test

## Data Availability

Data from the calculations is available in the supplementary files, and a spreadsheet with this calculated data is available: https://bit.ly/TestBasedCOVID19QuarantineData

There are no conflicts of interest to declare.

## Notes

### Competing Interest Statement

The authors have declared no competing interest.

### Funding Statement

No external funding was received.

## References

1. Bi Q, Wu Y, Mei S, Ye C, Zou X, Zhang Z, Liu X, Wei L, Truelove SA, Zhang T, Gao W. Epidemiology and transmission of COVID-19 in 391 cases and 1286 of their close contacts in Shenzhen, China: a retrospective cohort study. The Lancet Infectious Diseases. 2020 Apr 27.

2. Reich, N, Lauer, S, Li, X. “Determining Durations for Active Monitoring: Undetected Infections”. https://iddynamics.jhsph.edu/apps/shiny/activemonitr/. Accessed November 19, 2020.

3. Zhao Q, Ju N, Bacallado S, and Shah RD. BETS: The dangers of selection bias in early analyses of the coronavirus disease (COVID-19) pandemic. arXiv (2020).

4. Kucirka LM, Lauer SA, Laeyendecker O, Boon D, Lessler J. Variation in false-negative rate of reverse transcriptase polymerase chain reaction–based SARS-CoV-2 tests by time since exposure. Annals of Internal Medicine. 2020 May 23.

5. Washington State Coronavirus Response (COVID-19). https://coronavirus.wa.gov/what-you-need-know/safe-start/whats-open-each-phase. Accessed November 15, 2020.

