## Supplementary data Calculating Probability infected after quarantine with or without a negative test for "A Test-Based Strategy for Safely Shortening Quarantine for COVID-19"

Table S1: **Reich/Zhao estimates of number of undetected symptomatic COVID-19 infections per 10,000 monitored, assuming initial risk of 10%.**

| Duration, in days | Lower bound | Median | Upper bound |
|---|---|---|---|
| 1 | 903 | 933 | 959 |
| 2 | 763 | 811 | 859 |
| 3 | 622 | 678 | 733 |
| 4 | 499 | 552 | 610 |
| 5 | 391 | 443 | 497 |
| 6 | 304 | 350 | 399 |
| 7 | 232 | 273 | 317 |
| 8 | 174 | 212 | 252 |
| 9 | 130 | 163 | 198 |
| 10 | 96 | 125 | 155 |
| 11 | 71 | 95 | 121 |
| 12 | 52 | 72 | 94 |
| 13 | 38 | 54 | 74 |
| 14 | 27 | 41 | 57 |
| 15 | 20 | 31 | 44 |
| 16 | 14 | 23 | 34 |
| 17 | 10 | 17 | 26 |
| 18 | 7 | 13 | 20 |
| 19 | 5 | 10 | 16 |
| 20 | 4 | 7 | 12 |
| 21 | 3 | 5 | 9 |
| 22 | 2 | 4 | 7 |
| 23 | 1 | 3 | 6 |
| 24 | 1 | 2 | 4 |
| 25 | 1 | 2 | 3 |
| 26 | 0 | 1 | 3 |
| 27 | 0 | 1 | 2 |
| 28 | 0 | 1 | 1 |

**References:**

Reich, N, Lauer, S, Li, X. "Determining Durations for Active Monitoring: Undetected Infections". https://iddynamics.jhsph.edu/apps/shiny/activemonitr/. Accessed November 19, 2020.

Zhao Q, Ju N, Bacallado S, and Shah RD. BETS: The dangers of selection bias in early analyses of the coronavirus disease (COVID-19) pandemic. arXiv (2020).

Table S2: **Probability infected after quarantine with or without a negative test (following an initial 10% risk of infection)**

| Days since exposure | 1 | 2 | 3 | 4 | 5 | 6 | 7 | **8** | 9 | 10 | **11** | 12 | 13 | **14** | 15 | 16 | 17 | 18 | 19 | 20 | 21 |
|---|---|---|---|---|---|---|---|---|---|---|---|---|---|---|---|---|---|---|---|---|---|
| Probability PCR- given COVID+ (Kucirka) | 100% | 100% | 96% | 68% | 38% | 25% | 21% | 20% | 21% | 24% | 27% | 31% | 35% | 39% | 44% | 48% | 52% | 56% | 60% | 64% | 68% |
| Proportion of infections showing symptoms (Bi) | 1% | 9% | 23% | 39% | 52% | 64% | 72% | 79% | 84% | 87% | 90% | 92% | 94% | 95% | 96% | 97% | 98% | 99% | 99% | 99% | 99% |
| Undetected symptomatic infections rate (Reich, Zhao @ 10%) | 9.3% | 8.1% | 6.8% | 5.5% | 4.4% | 3.5% | 2.7% | 2.1% | 1.6% | 1.2% | 1.0% | 0.7% | 0.5% | 0.4% | 0.3% | 0.2% | 0.2% | 0.1% | 0.1% | 0.1% | 0.1% |
| Undetected symptomatic infections rate (1-Bi * 10%) | 9.9% | 9.1% | 7.7% | 6.1% | 4.8% | 3.6% | 2.8% | 2.1% | 1.6% | 1.3% | 1.0% | 0.8% | 0.6% | 0.5% | 0.4% | 0.3% | 0.2% | 0.2% | 0.1% | 0.1% | 0.1% |
| Probability infected after quarantine only | 9.6% | 8.6% | 7.2% | 5.8% | 4.6% | 3.5% | 2.8% | 2.1% | 1.6% | 1.3% | 1.0% | 0.8% | 0.6% | **0.5%** | 0.4% | 0.3% | 0.2% | 0.1% | 0.1% | 0.1% | 0.1% |
| Probability infected after quarantine followed by a negative test | 9.6% | 8.6% | 7.0% | 4.1% | 1.8% | 0.9% | 0.6% | **0.4%** | 0.3% | 0.3% | **0.3%** | 0.2% | 0.2% | **0.2%** | 0.2% | 0.1% | 0.1% | 0.1% | 0.1% | 0.1% | 0.1% |

**References:**


Reich, N, Lauer, S, Li, X. "Determining Durations for Active Monitoring: Undetected Infections". https://iddynamics.jhsph.edu/apps/shiny/activemonitr/. Accessed November 19, 2020.

Zhao Q, Ju N, Bacallado S, and Shah RD. BETS: The dangers of selection bias in early analyses of the coronavirus disease (COVID-19) pandemic. arXiv (2020).

Bi Q, Wu Y, Mei S, Ye C, Zou X, Zhang Z, Liu X, Wei L, Truelove SA, Zhang T, Gao W. Epidemiology and transmission of COVID-19 in 391 cases and 1286 of their close contacts in Shenzhen, China: a retrospective cohort study. The Lancet Infectious Diseases. 2020 Apr 27.

Kucirka LM, Lauer SA, Laeyendecker O, Boon D, Lessler J. Variation in false-negative rate of reverse transcriptase polymerase chain reaction–based SARS-CoV-2 tests by time since exposure. Annals of Internal Medicine. 2020 May 23.


A spreadsheet with this calculated data is available: https://bit.ly/TestBasedCOVID19QuarantineData
